# MpoxPlex: a high-throughput and versatile multiplexed immunoassay for assessing and discriminating between IgG responses to Mpox infection and vaccination

**DOI:** 10.1101/2024.06.03.24308186

**Authors:** Scott Jones, Bethany Hicks, Helen Callaby, Daniel Bailey, Claire Gordon, Tommy Rampling, Catherine Houlihan, Rachael Jones, Marcus Pond, Ravi Mehta, Deborah Wright, Clarissa Oeser, Simon Tonge, Ezra Linley, Cathy Rowe, Bassam Hallis, Ashley Otter

## Abstract

The summer of 2022 saw the first global outbreak of Mpox disease (formerly ‘monkeypox’), primarily within gay, bisexual, and other men who have sex with men (GBMSM). In response, public health agencies in the UK have offered smallpox vaccines to those individuals deemed at highest risk of infection. With Mpox cases still being detected globally, novel tools are required to aid with diagnosis, serosurveillance and the evaluation of immune responses following infection and vaccination. Here, we describe the development of a multiplexed immunoassay that is able to measure IgG responses to twelve immunogenic Orthopoxvirus proteins concurrently and distinguish between responses to infection and vaccination.

Using the Luminex platform, antibody responses to vaccinia virus (VACV) proteins B5, A27, A33 and Monkeypox virus (MPXV) proteins E8, B6, B2, M1, A27, A35, H3, A29, A5 were assessed in serum from individuals post-MPXV infection (n=24) and post-vaccination (n=75) with modified vaccinia virus Ankara-Bavarian Nordic (MVA-BN, “IMVANEX”). Negative sera (n=435) were run alongside to assess appropriate assay cut-offs and characteristics.

Using the results from a combination of eight of the twelve proteins within the immunoassay we were able to classify samples as either post-vaccination or infection, from negative samples with a sensitivity of 98.39% (9.72-99.22%) and specificity of 95.24% (86.91-98.70%). IgG responses to VACV A27, MPXV A29 and MPXV A5 provided little diagnostic advantage. IgG responses to the MPXV protein A27 were able to distinguish post-MPXV infection from negative and post-vaccination samples with a sensitivity of 87.5% (69.00-95.66%) and specificity of 96.84% (94.84-98.07%).

There is an ongoing need to utilise Mpox serology to conduct disease surveillance, assess the efficacy of current and new vaccine candidates, and further understand immune responses to Mpox infection. We believe this assay will provide substantial insight into the current global outbreak of Mpox, with additional benefits over current serological assays.

## Introduction

First described in 1959 after an outbreak in two Cynomolgus monkey colonies, monkeypox virus (MPXV) is an Orthopoxvirus virus capable of causing Mpox disease (formerly ‘Monkeypox’) in humans [1–3]. Mpox disease was identified in humans during the latter stages of Smallpox eradication when a handful of individuals living in areas of West Africa, considered free of the variola virus, presented with symptoms akin to smallpox [4–6]. Isolation of the causative agents were distinct from variola virus but indistinguishable from previous MPXV isolates. Until recently, reports of human-to-human transmission were limited and MPXV was considered to primarily be a zoonosis with many mammalian species considered reservoirs [7].

Before the 2000s, confirmed cases of Mpox disease were rare (though increasing) and limited to countries of Western and Central Africa, with the vast majority occurring in the Democratic Republic of the Congo (DRC) [8]. Following the sudden re-emergence of Mpox disease in Nigeria in 2017 and increasing cases in the DRC, a small number of imported cases were reported in the United Kingdom, Israel, and Singapore [8–10]. Outside of endemic regions, limited and local transmission of MPXV had only ever been reported in the United States (with transmission from infected prairie dogs to humans) and the UK (human to human between an infected individual and a healthcare worker) prior to 2022 [11, 12]. The notably milder disease produced by the virus responsible for the outbreak in the United States resulted in the identification of two distinct MPXV clades, designated Clade I (formerly known as the Central African Clade or Congo Basin Clade) and Clade II (formerly known as the West African Clade)[13]. In general, Clade I has been observed to produce more severe disease, longer viraemia, higher rates of fatality and has an increased chance of human-to-human transmission [13]. Clade II is considered less virulent with a fatality rate of <0.2%, significantly lower than the 10% case fatality rate observed with Clade I MPXV [14, 15].

Since May 2022, an ongoing global outbreak of Clade IIb Mpox has result in an unprecedented 93,030 confirmed cases in 117 countries, many of which have never previously reported the disease [16]. Worldwide, Mpox cases peaked in mid-2022 with over 30,000 confirmed in August alone. Most cases were present in the WHO region of the Americas and European region. Since the peak of the outbreak in August 2022, cases numbers have fallen and have remained low in the Americas and Europe. More recently in mid-late 2023, case numbers have been highest in the African, South-East Asian and Western Pacific Regions[16–20]. The predominant mode of transmission within the recent outbreak has primarily been through sexual contact and has principally affected gay, bisexual and other men who have sex with men (GBMSM) [21]. As such, public health interventions have focussed their efforts on these individuals by raising awareness and offering testing and vaccination in areas of high transmission. At present, there is a large Clade I outbreak in the DRC and investigations have shown similar routes of transmission [22].

There are currently no licensed vaccines against Mpox disease. However, numerous animal challenge studies and retrospective analysis of vaccination status in Mpox outbreaks have shown Smallpox vaccines to be protective against disease, with an estimated efficacy of approximately 79-93% [23–27]. It has also been suggested that Mpox disease severity and outcome is reduced in those previously vaccinated against smallpox [28, 29]. Nevertheless, the longevity of protection provided by smallpox vaccination against Mpox disease is unknown. Current smallpox vaccines include live, attenuated vaccinia virus strains derived from the Lister or modified vaccinia Ankara (MVA) [30, 31]. In many countries, including the UK, routine Smallpox vaccination was discontinued following the eradication of the disease in the 1970-80s, which has led to the hypothesis of populations increasingly susceptible to Orthopoxvirus outbreaks such as Mpox [32]. The next generation of Orthopoxvirus vaccines include subunit and mRNA vaccines, with some specifically for Mpox [33–37]. Since the May 2022 Mpox outbreak, Smallpox vaccination has been recommended to those deemed at high risk of infection [38].

MPXV and Vaccinia virus (VACV) express over 200 proteins. Immune responses to infection and vaccination are complex [39, 40]. A number of studies prior to the 2022 outbreak identified core immunodominant antigens in response to infection and vaccination [41–44]. The antigens VACV A33, L1, B5, A56 and H3 (homologues of MPXV A35, M1, B6, B2 and H3, respectively) have repeatedly been acknowledged to induce robust antibody responses in the host and to play a significant role in immunity [37, 41, 43–47]. Antibodies to these antigens have been shown to be sufficient for protection against severe disease in animal challenge models and form the basis for many subunit vaccines [36, 48–52]. We recently characterised the antibody responses in individuals infected with Clade II(b) and vaccinated with MVA-Bavarian Nordic (IMVANEX) highlighting additional immunodominant antigens, such as MPXV A5, A27, A29 and E8 [45]. We also demonstrated the similarity between antibody response profiles to vaccination and infections with exception of MPXV A27, which, crucially, is unique to convalescent Mpox sera. However, this characterisation of immune responses was time and resource intensive, requiring individual antigen ELISAs, which would be inappropriate in settings requiring high throughput testing, such as for vaccine testing and serosurveillance studies. Transfer of these assays to platforms capable of multiplexing, such as the Luminex system, addresses this issue whilst maintaining the volume of data produced.

As cases continue to be detected, serological assays are needed for use in diagnosis, serosurveillance, and the understanding of immune responses to infection and vaccination. Here, we describe the development of an assay capable of measuring IgG responses to twelve Orthopoxvirus proteins simultaneously, for use in MPXV serosurveillance and immunological studies of vaccination and infection.

## Materials and Methods

### Samples and Ethics

Post-MPXV Clade II(b) infection sera from the 2022 outbreak were obtained from the Rare and Imported Pathogens Laboratory (RIPL, UKHSA), Imperial College Healthcare NHS Trust, and Chelsea and Westminster Hospital NHS Foundation Trust (n=22; median of 81 days post infection). Post-MPXV Clade II(a) infection sera from 2018-2019 were from imported cases obtained from RIPL, UKHSA (n=2; median of 7 days post infection). Post-MPXV infection sera were from individuals with a median age of 37 years. NHS Research Ethics Committees (REC) granted approval for sampling from previous MPXV-infected individuals under reference 22/HRA/3321, with residual and fully anonymised serum from individuals with prior MPXV infection sourced from diagnostic laboratories for surveillance, assay performance assessment, validation, and public health monitoring.

Paediatric negative samples were obtained from the UKHSA Seroepidemiology Unit, Manchester (n=224; median age of 3 years). Negative confounder serum samples PCR-positive for cytomegalovirus (CMV; n=52; median age of 34 years), Epstein Barr virus (EBV; n = 34; median age of 25 years) and Varicella zoster virus (VZV; n = 44; median age of 32 years) were used to determine assay specificity. Serum samples positive for Rheumatoid factor (RF; n = 63; median age of 15 years) were also used to assess assay characteristics. Pre-IMVANEX vaccination sera were obtained from UKHSA, Porton Down (n=18; median age of 27 years).

Post-IMVANEX vaccination sera were obtained at various timepoints post vaccination from UKHSA, Porton Down (n=75; median age of 26 years). Samples obtained from IMVANEX Smallpox-vaccinated individuals were obtained through written and informed consent through UKHSA Research and Ethics Committee (“REGG”) for assay validation. Further details of convalescent serum from Mpox PCR-confirmed cases, serum from smallpox-vaccinated individuals, negative samples and confounder samples are as described by Otter, Jones and Hicks et al., 2023 and in **Supplementary Table S1** [45].

### MPXV and VACV Antigens

Recombinant MPXV proteins A27 (product code PXP6054), B6 (PXP6031) and E8 (PXP6055) were purchased from ProteoGenix (France), A29 (40891V08E), A35 (40886V08H), H3 (40893V08H1) and M1 (40904V07H) from SinoBiological (Germany), B2 (ABX620113) from Abbexa (UK), and A5 (REC32033) from Native Antigen Company (UK). Recombinant VACV Copenhagen proteins A27 (40897V07H), A33 (40896V07E) and B5 (40900V08H) were purchased from SinoBiological. All lyophilised proteins were reconstituted as directed by the manufacturers and stored at -80°C prior to use.

### Multiplexed Mpox Assay

#### Microsphere Coupling

MPXV and VACV antigens were coupled to Luminex MagPlex® microspheres (DiaSorin, Italy) as per the manufacturer’s recommendations. Reagents used for microsphere coupling were obtained from the Luminex xMAP® antibody coupling kit (product code 40-50016) from DiaSorin. Each MPXV and VACV antigen was coupled to a different bead region, utilising bead regions 12-15, 18, 20-22, 25, 35, 37 and 38.

Microsphere stocks were first resuspended by vortexing and sonicating for 30 seconds and 240µl was aliquoted into a 2ml low binding tube. Tubes were then placed into a magnetic rack separator (Invitrogen, USA, product code CS15000) for 60 seconds and the supernatant was aspirated whilst still in the separator. Tubes were removed from the rack and the microspheres were resuspended in 240µl ddH_2_O, vortexed and sonicated as before. Tubes were placed back into the magnetic rack for 60 seconds and the supernatant was aspirated before being removed and resuspended in 80µl activation buffer, vortexed and sonicated as before. 10µl Sulfo-NHS solution and 10µl 40mg/ml ECD solution was added to the microspheres and tubes were vortexed and sonicated. Tubes were then wrapped in foil and placed into a tube rotator at 10 RPM for 20 minutes at ambient temperature. Tubes were placed back into the magnetic rack for 60 seconds and the supernatant was aspirated before being removed and resuspended in 250µl activation buffer, vortexed and sonicated as before. This step was repeated to fully remove the Sulfo-NHS and ECD from the microspheres. Tubes were placed back into the magnetic rack for 60 seconds and the supernatant was aspirated before being removed and resuspended in 500µl of antigen (diluted to 12µg/ml in activation buffer) vortexed and sonicated. Tubes were then wrapped in foil and placed into a tube rotator at 10 RPM for 120 minutes at ambient temperature. Tubes were placed back into the magnetic rack for 60 seconds and the supernatant was aspirated before being removed and resuspended in 500µl PBS with 0.05% Tween20 (ThermoFisher, USA), 0.1% BSA (SLS, UK) and 0.02% sodium azide (VWR, UK), vortexed and sonicated as before. This step was repeated with 1000µl PBS with 0.05% Tween20, 0.1% BSA and 0.02% sodium azide to fully remove unbound antigen from the microspheres. Tubes were placed back into the magnetic rack for 60 seconds and the supernatant was aspirated before being removed and resuspended 1000ul PBS-1% BSA, 0.05% sodium azide and stored at 4°C until required. The concentration of antigen-coupled microspheres was measured using a TC20™ automated cell counter (Bio-Rad, UK).

#### Sample Testing

Microspheres coupled to each MPXV and VACV antigen were diluted to 50 microspheres/µl with PBS 0.05% Tween20 1% BSA in the same tube and mixed well using a vortex and sonicator. Samples were diluted 1:25 with PBS 0.05% Tween20 1% BSA. In a low binding 96-well microplate (Corning, USA, product code 3474), 50µl/well of both the diluted microsphere panel and samples were combined, mixed, and incubated at ambient for 30 minutes with shaking at 700 RPM. During incubation steps, plates were sealed and covered in foil to minimise photobleaching. Plates were then washed three times with 200µl/well PBS 0.05%Tween20 using a 405 TS BioTek microplate washer (Agilent, USA) fitted with a microplate magnet (product code 7103016) to avoid microsphere loss in washing stages. Microspheres were resuspended in 100µl/well PE-conjugated mouse anti-human IgG antibody (AbCam, UK, product code ab99761), diluted 1:500 in PBS 0.05%Tween20, before being incubated at ambient for 30 minutes with shaking at 700 RPM. Plates were then washed as before, and microspheres were resuspended in 100µl/well PBS 0.05%Tween20 and mixed on a microplate shaker for five minutes at 700RPM before reading. Plates were read using a Luminex xMAP INTELLIFLEX® system (DiaSorin) at a low PMT setting with a minimum bead count of 100/well per region.

#### Singleplex Mpox ELISAs

Singleplex Mpox ELISAs were run as described previously [45]. Briefly, samples, diluted 1:200 in Superblock (ThermoFisher, product code 37515), were incubated for 1 hour at 37°C in high-binding, flat-bottom, 96-well microplates (Corning, USA, product code 9018) coated overnight at 4°C with 100µl/well of 0.1µg/ml antigen or PBS (Gibco, USA). Bound, antigen-specific IgG were detected by adding 100µl/well HRP-conjugated goat anti-human IgG preabsorbed H+L antibody (AbCam, UK, product code AB99761), diluted 1:8000 in Superblock. Plates were developed for 15 minutes at 37°C by adding 50 µl/well 1-Step™ Ultra TMB-ELISA Substrate Solution (ThermoFisher, product code 34029), stopped by adding 50 µl/well KPL TMB Stop Solution (Seracare, USA, product code 5150-0021) and read at an absorbance of 450nm using an Infinite F50 plate reader (Tecan, Switzerland). Data obtained by the singleplex Mpox ELISAs were analysed as described by Otter, Jones and Hicks et al., 2023 [45].

### Data Analysis

Data handling and analyses, and graph and table generation were performed using either Excel (version 2208; Microsoft, USA) and GraphPad Prism (version 9.2.0; GraphPad, USA). Sample median fluorescence intensities (MFIs) were generated using IFLEX software (version 2.1; Diasorin).

#### Individual Antigen Assay Cut-offs

MFI cut-offs were calculated for each antigen by fitting receiver operating characteristic (ROC) curves to the MFIs of negative samples (negative confounders: n=193; paediatric negatives: n=224 and pre-IMVANEX vaccination: n=18) versus positive samples (post-MPXV infection: n=24 and or post-IMVANEX vaccination timepoints between 14 to 144 days post-two doses: n=39) (**Table 1**). Cut-offs were selected based on the highest sum of sensitivity and specificity.

**Table 1.** Summary of ROC Curves and Cut-offs for Each Antigen (Positive Versus Negative Samples) Summary characteristics of curves and optimal cut-offs generated by comparing the MFIs of all positive samples (n=63) and all negative samples (n=435) to 12 MPXV & VACV antigens by ROC analysis. Cut-offs were selected based on the highest sum of sensitivity and specificity.

| ROC curve characteristics |  |  |  |  | MFI cut-off characteristics |  |  |  |  |
| --- | --- | --- | --- | --- | --- | --- | --- | --- | --- |
| Antigen | Area | Std. Error | 95% confidence interval | P value | MFI | Sensitivity% | 95% CI | Specificity% | 95% CI |
| VACV B5 | 0.980 | 0.01097 | 0.9587 to 1.000 | <0.0001 | > 87.41 | 94.94 | 92.46% to 96.64% | 95.24 | 86.91% to 98.70% |
| MPXV E8 | 0.980 | 0.01323 | 0.9537 to 1.000 | <0.0001 | > 101.3 | 96.55 | 94.39% to 97.90% | 95.24 | 86.91% to 98.70% |
| MPXV B6 | 0.978 | 0.0125 | 0.9533 to 1.000 | <0.0001 | > 64.35 | 94.71 | 92.19% to 96.45% | 95.24 | 86.91% to 98.70% |
| MPXV B2 | 0.977 | 0.01362 | 0.9507 to 1.000 | <0.0001 | > 41.24 | 93.1 | 90.33% to 95.13% | 96.83 | 89.14% to 99.44% |
| VACV A33 | 0.960 | 0.0163 | 0.9281 to 0.9920 | <0.0001 | > 294.9 | 92.41 | 89.54% to 94.55% | 93.65 | 84.78% to 97.50% |
| MPXV M1 | 0.959 | 0.01187 | 0.9357 to 0.9822 | <0.0001 | > 31.16 | 90.11 | 86.95% to 92.58% | 92.06 | 82.73% to 96.56% |
| MPXV A35 | 0.952 | 0.0165 | 0.9194 to 0.9841 | <0.0001 | > 136.5 | 91.26 | 88.24% to 93.57% | 87.3 | 76.89% to 93.42% |
| MPXV H3 | 0.943 | 0.01871 | 0.9066 to 0.9799 | <0.0001 | > 296.4 | 91.72 | 88.76% to 93.96% | 87.3 | 76.89% to 93.42% |
| MPXV A29 | 0.720 | 0.03132 | 0.6586 to 0.7814 | <0.0001 | > 30.17 | 73.79 | 69.47% to 77.70% | 63.49 | 51.15% to 74.28% |
| MPXV A5 | 0.711 | 0.03358 | 0.6450 to 0.7766 | <0.0001 | > 156.4 | 63.91 | 59.29% to 68.28% | 71.43 | 59.30% to 81.10% |
| MPXV A27 | 0.699 | 0.03849 | 0.6231 to 0.7739 | <0.0001 | > 259.5 | 86.67 | 83.15% to 89.54% | 46.03 | 34.31% to 58.21% |
| VACV A27 | 0.646 | 0.04079 | 0.5655 to 0.7255 | 0.0002 | > 281.4 | 85.29 | 81.65% to 88.31% | 44.44 | 32.85% to 56.68% |

#### Screening Assay Cut off

For each sample, the number of antigens with MFIs above the cut-off was calculated. To assess which combination of antigens were most well suited to differentiate negative and positive samples, the ‘antigen positivity’ scores were compared by fitting ROC curves to the antigen with the highest area under the curve followed by the next highest antigen stepwise until the scores of all twelve antigens were combined (**Table 2** and **Supplementary Figure S1**). The order of antigens was VACV B5 followed by MPXV E8, B6, B2, VACV A33, MPXV M1, A35, H3, A29, A5, A27 and VACV A27. The antigen positivity score cut-off was selected based on the highest sum of sensitivity and specificity.

**Table 2.**
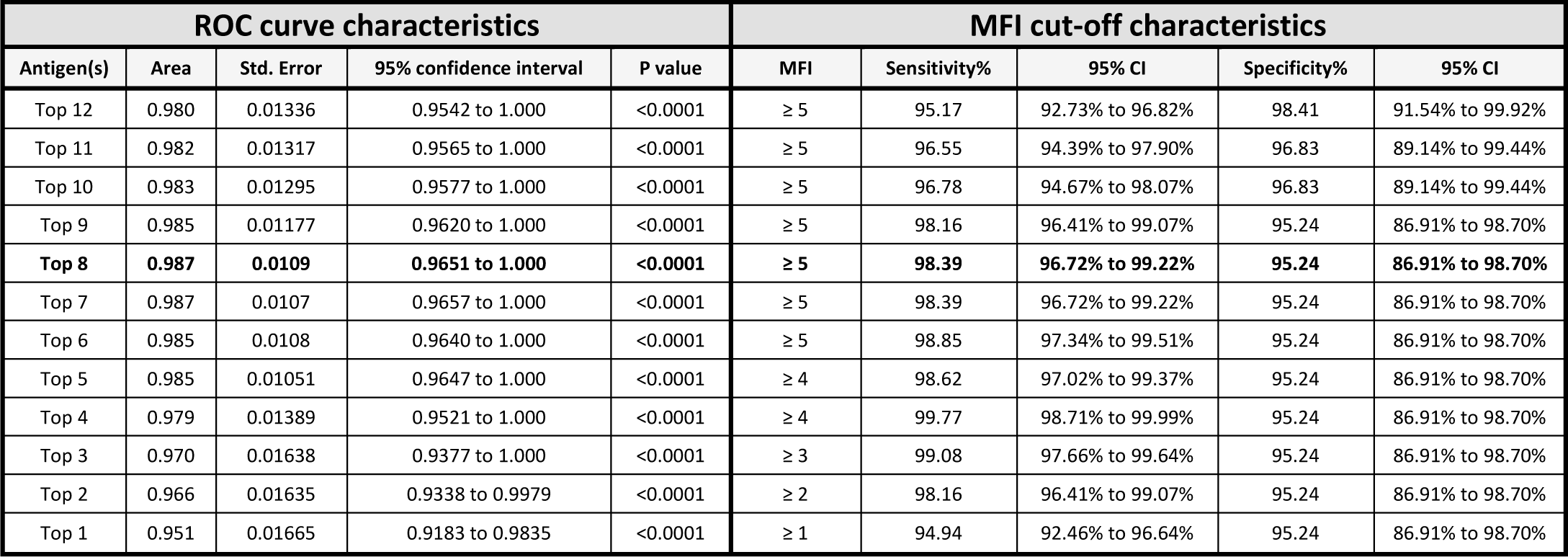
Summary of ROC Curves and Positive Score Cut-offs for Antigen Combinations (Positive Versus Negative Samples) Summary characteristics of curves and optimal cut-offs generated by comparing the positivity scores of all positive samples (n=63) and all negative samples (n=435) to 12 MPXV & VACV antigens by ROC analysis. Positivity scores were calculated based on how many antigen cut-offs a sample was above (one for each antigen). The order of antigens was the following: VACV B5 (top), MPXV E8, B6, B2, VACV A33, MPXV M1, A35, H3, A29, A5, A27 and VACV A27 (bottom). For example, Top 1 includes VACV B5 only, Top 2 includes VACV B5 and MPXV E8 and so on. Cut-offs were selected based on the highest sum of sensitivity and specificity.

#### Differential Assay Cut-off

The ability of the antigens not included in the screening assay (MPXV A29, A5, A27 and VACV A27) to differentiate post-MPXV infection samples from post-IMVANEX vaccination samples was quantified by fitting ROC curves to the MFIs for each antigen for all negative plus post-IMVANEX vaccination samples versus post-MPXV infection samples (**Table 3**). Cut-offs for each antigen were selected based on the highest sum of sensitivity and specificity.

**Table 3.**
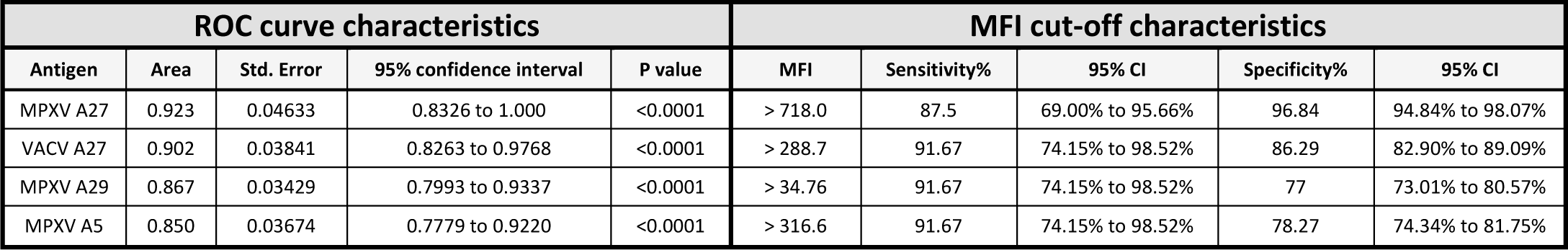
Summary of ROC Curves and Positive Score Cut-offs for Select Antigens (Post-MPXV Infection Versus Negative and Post-IMVANEX Samples) Summary characteristics of curves and optimal cut-offs generated by comparing the MFIs of post-MPXV infection (n=24) with all negative samples (n=435) and post-IMVANEX vaccination (n=39, 14 to 144 days post-two doses) to MPXV A27, A29, A5 and VACV A27 antigens by ROC analysis. Cut-offs were selected based on the highest sum of sensitivity and specificity.

## Results

### Assessment of IgG Responses to 12 MPXV and VACV Antigens

The MFIs of post-MPXV infection (n=24), post-IMVANEX vaccination (timepoints between 14 to 144 days post-two doses; n=39) and negative samples (pre-IMVANEX vaccination: n=18; paediatric negatives: n=224 and negative confounders: n=193) to 12 MPXV and VACV antigens were compared (**Figure 1** and **Table S3**). Post-MPXV infection and post-IMVANEX vaccination samples were significantly higher than the negatives for MPXV antigens B6, E8, A35, B2, M1 and H3, and VACV antigens A33 and B5. In comparison, the MFIs for MPXV antigens A5, A27 and A29 and VACV A27 were only significantly higher than the negatives for post-MPXV infection samples.

**Figure 1.**
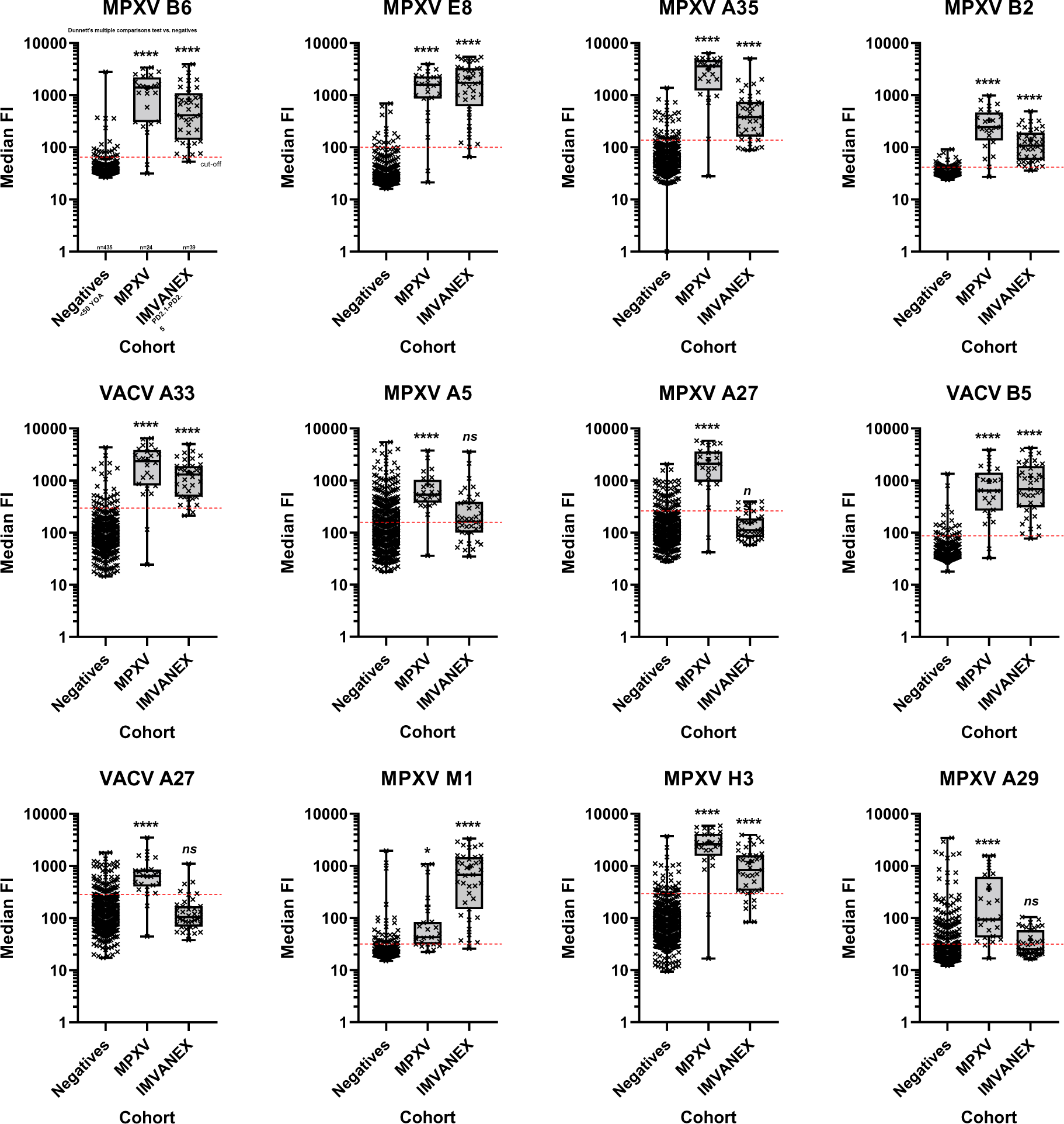
Antigen MFIs of Negative, Post-MPXV Infection and Post-IMVANEX Vaccination Samples. Sample MFIs run on the multiplexed Mpox assay for each antigen. Each graph represents the results for an individual antigen. Each cross represents the MFI for one sample. From top to bottom, the horizonal lines of the box and whisker plots represent the maximum, 75^th^ percentile, median, 25^th^ percentile and minimum MFIs. Red dotted line represents MFI cut-offs as calculated in **Table 1**. Significance compared to the negative samples was calculated using Dunnette’s multiple comparisons test (ns = not significant; * = <0.05; **** = <0.0001). The plus symbol represents the mean. Negative samples (n= 435); post-MPXV infection (n=24); post-IMVANEX vaccination (n=39, 14 to 144 days post-two doses).

### Analysis of Multiplexed Mpox Assay Results for Diagnosis

When comparing the MFIs of all negative samples versus positive (post-MPXV and vaccination) samples, the twelve antigens tested varied in their sensitivities and specificities for detecting antibodies induced by vaccination or infection, with ROC area under the curves ranging from 0.646 for VACV A27 to 0.980 for VACV B5 (**Table 1**).

ROC curves were also generated comparing the MFIs of each of the sample groups separately (**Figure 2**). For negative samples versus post-MPXV infection samples, area under the curves for all antigens were comparable ranging from 0.854 for MPXV A5 to 0.962 for B2 (**Figure 2A**). For negative samples versus post-IMVANEX vaccination samples, area under the curves for MPXV E8, B6, B2, M1, A35 and H3, and VACV B5 and A33 antigens were similar ranging between 0.942 and 0.994 (**Figure 2B**). Notably, area under the curves for MPXV A29, A5 and A27, and VACV A27 antigens were much lower ranging between 0.511 and 0.629. The ROC curves for post-MPXV infection samples versus post-IMVANEX vaccination samples were more variable (**Figure 2C**). Area under the curves for MPXV A27 and VACV A27 were the highest at 0.922 and 0.9145, respectively. Conversely, area under the curves for MPXV E8 and VACV B5 were the lowest at 0.5769 and 0.5406, respectively

**Figure 2.**
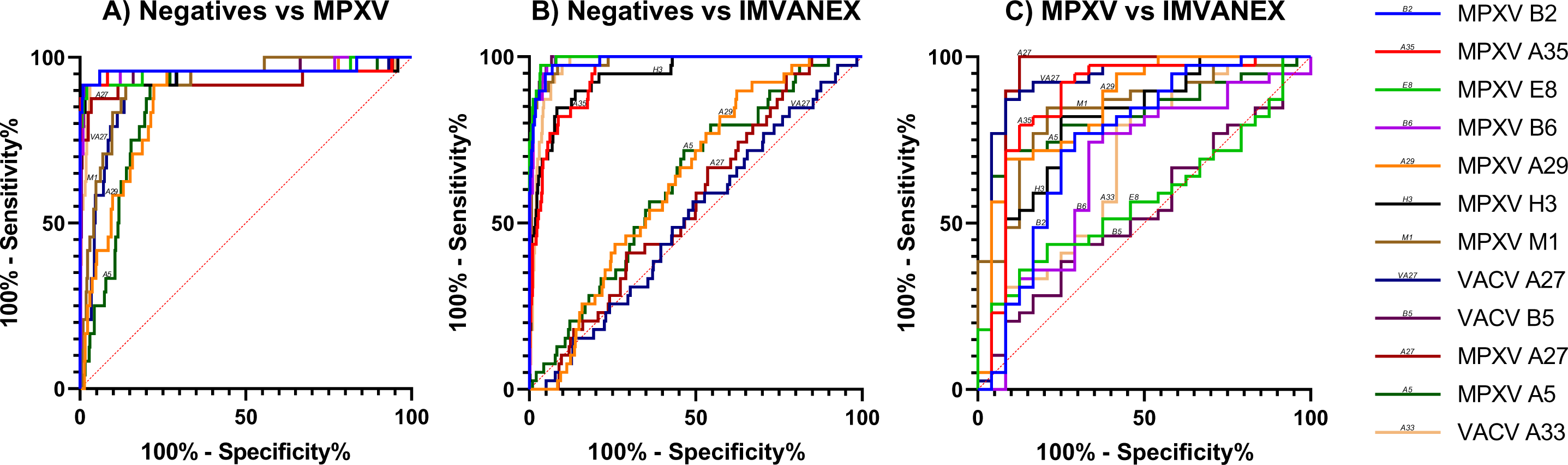
Individual Antigen ROC Curves (Negative Samples Versus Post-MPXV Infection or Post-IMVANEX Vaccination Samples) ROC curves of the MFIs of negative samples versus post-MPXV infection samples (**A**), negative samples (n=435) versus post-IMVANEX vaccination samples (n=39, 14 to 144 days post-two doses) (**B**) or post-MPXV infection (n=24) versus post-IMVANEX vaccination samples (**C**) of each of the 12 MPXV and VACV antigens tested. Each solid line represents the ROC curve for an individual antigen. Dotted red line represents the random classifier (i.e., sensitivity = specificity).

By comparing the ability of different antigen combinations to differentiate negative and positive samples as described in the methods, the top eight antigens (MPXV E8, B6, B2, M1, A35 and H3, and VACV B5 and A33 antigens) was determined optimal for classifying negative and positives samples with an area under the curve of 0.987 and, using an ‘antigen positivity’ score cut-off of greater than 4, a sensitivity of 98.39% and specificity of 95.24% (**Table 2** and **Supplementary Figure S1**). Including more or fewer antigens resulted in a poorer performance.

The ability of the remaining four antigens (MPXV A29, A5, A27, and VACV A27) to identify post-MPXV infection samples was assessed as described in the methods (**Table 3**). Area under the curves for all antigens were similar (ranging between 0.810 and 0.923) with MPXV A27 performing the best, followed by VACV A27, MPXV A29 and MPXV A5. From this, it was concluded that MPXV A27 alone was optimal for distinguishing post-MPXV infection samples from all other samples with 87.5% sensitivity (69.00 - 95.66) and 96.84% (94.84 - 98.07) specificity.

To summarise, we have developed a method whereby if a sample has MFIs above the cut-offs for at least five of the eight antigens it is considered positive for Orthopox-specific antibodies, induced by infection or vaccination (screening assay). If the sample is above the MFI cut-off for MPXV A27, it is then classified as post-MPXV infection, whilst negative result for A27 is classified as post-IMVANEX vaccination (differential assay).

### Correlation of Multiplexed Mpox Assay and Singleplex Mpox ELISAs

The MFIs of post-MPXV infection (n=24), paediatric negatives (n=17), pre-IMVANEX vaccination (n=16), and post-IMVANEX vaccination (timepoints between 14 to 144 days post-dose (PD) 2; n=35) samples with corresponding ELISA data were compared for the twelve antigens assessed by the multiplexed Mpox assay (**Figure 3** and **Table S1**). The line of best-fit for each antigen was calculated using a hyperbola non-linear regression model. The results from both assays showed a good correlation for antigens VACV B5, A33 and A27, and MPXV A35, B6, B2, A27, E8, and M1 with R-squared values ranging from 0.513 to 0.793. Antigens MPXV A5, A29 and H3 showed little-to-no correlation with R-squared values of 0.1642, 0.030 and 0.020, respectively.

**Figure 3.**
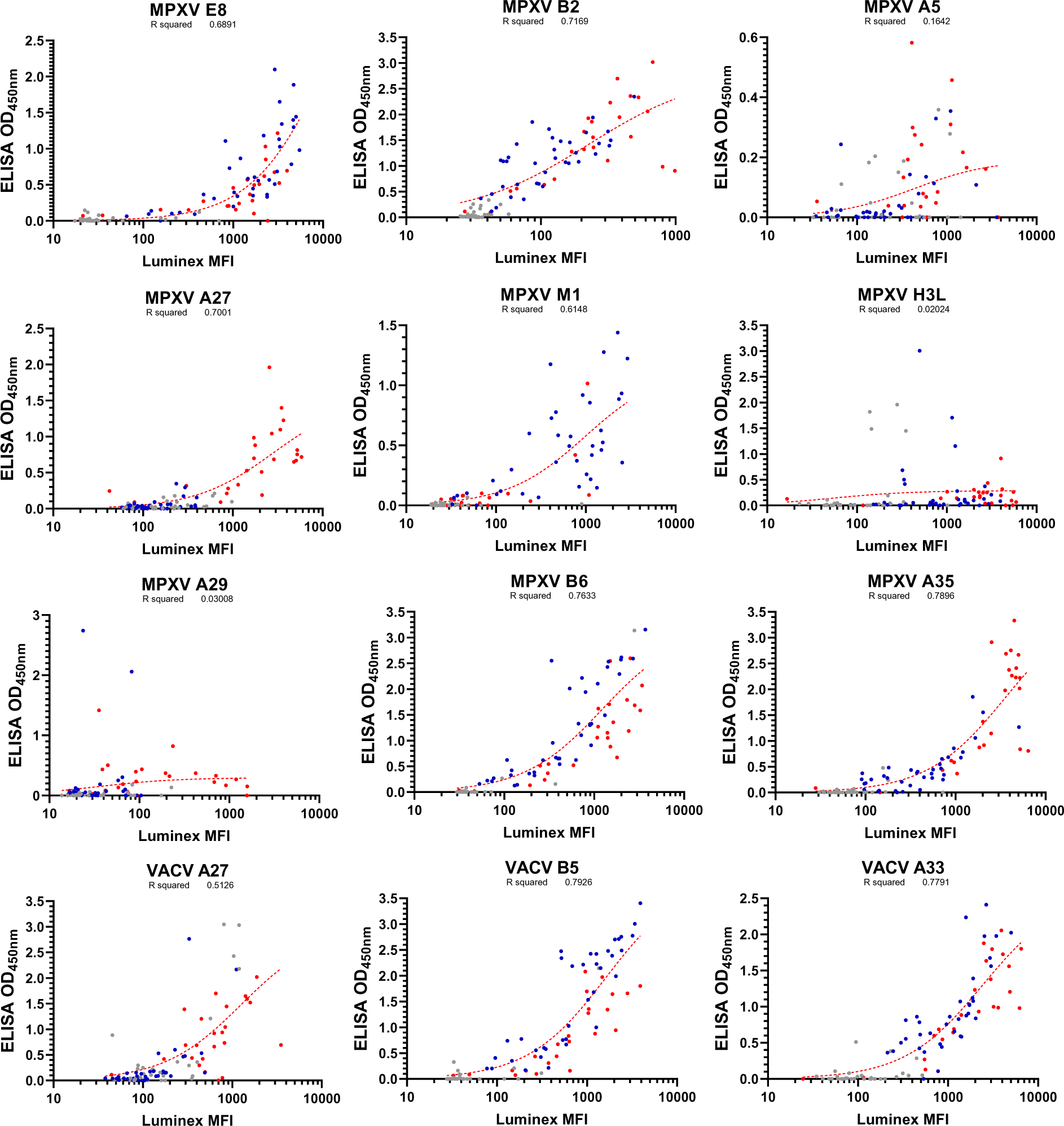
Correlation of Multiplexed Mpox Assay and Singleplex Mpox ELISAs. Samples were run on both the multiplexed Mpox Luminex assay and singleplex Mpox ELISAs against 12 MPXV and VACV antigens and results were correlated. The lines of best fit were calculated using a hyperbola non-linear regression model (dotted, red line). Each circle represents the ELISA (OD_450nm_) and Luminex (MFI) results for one sample. Post-MPXV infection samples (n=24, red circles); paediatric negative samples (n=17, grey circles); pre-IMVANEX vaccination samples (n=16, grey circles); post-IMVANEX vaccination samples (n=35, timepoints between 14 to 144 days post two doses; blue circles).

### Differential IgG Responses of Post-MPXV Infection or IMVANEX Vaccination to MPXV and VACV Homologous Antigens

The ratios of the MFIs for MPXV antigens B6, A35 and A29 and their VACV homologues (B5, A33, and A27, respectively) were compared for post-MPXV infection (n=24) and post-IMVANEX vaccination (timepoints between 14 to 144 days PD2; n=39) samples (**Figure 4**). By unpaired T-test, the ratios of MPXV:VACV MFIs were significantly higher in samples taken post-MPXV infection compared to samples taken post-IMVANEX vaccination. The mean MPXV:VACV MFI ratios for post-MPXV infection samples were 1.596, 1.399 and 2.125 for antigens MPXV B6:VACV B5, MPXV A35:VACV A33 and MPXV A29:VACV A27, respectively. In comparison, the mean MPXV:VACV MFI ratios for post-IMVANEX vaccination samples were 0.664, 0.389 and 0.351.

**Figure 4.**
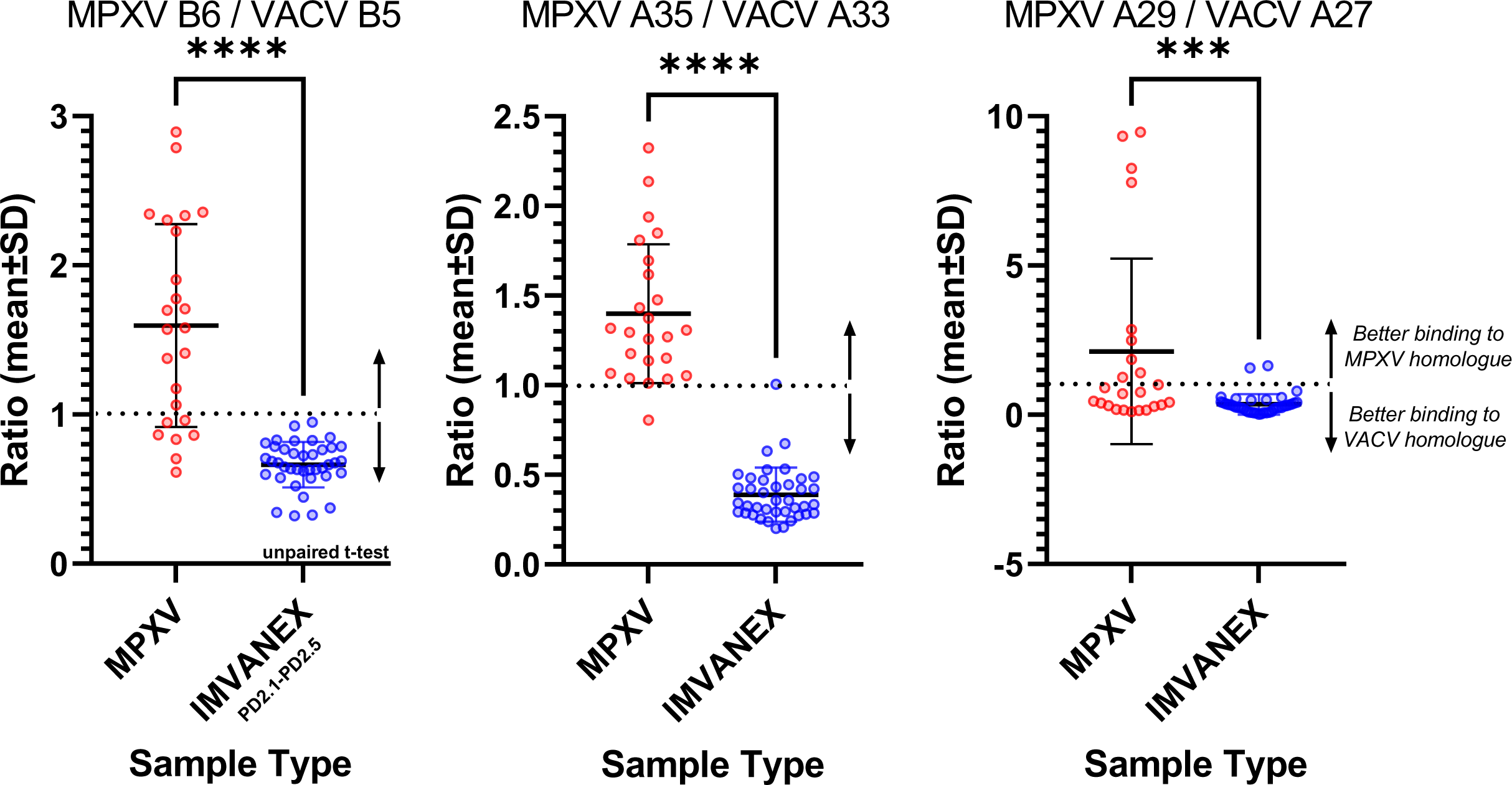
Comparison of Responses to MPXV and VACV Homologues in MPXV Infection and IMVANEX Immunisation. The ratios of the MFIs for MPXV and VACV antigen homologues MPXV B6 and VACV B5, MPXV A35 and VACV A33, and MPXV A29 and VACV A27 for post-MPXV infection and post-IMVANEX vaccination samples were compared. The significant difference between the two groups was assessed using an unpaired t-test (*** = <0.001; **** = <0.0001). Post-MPXV infection samples (n=24, red circles); post-IMVANEX vaccination samples (n=39, timepoints between 14 to 144 days post two doses; blue circles). Each circle represents the ratio of a single sample. Black lines represent the mean ratio ± standard deviation.

### Longitudinal IgG Responses to 12 MPXV and VACV Antigens Post-IMVANEX Vaccination

The responses of samples post one (PD1) and two (PD2) doses of IMVANEX vaccine, taken at multiple different timepoints, to all 12 antigens were assessed using the described assays (**Figure 5**). Vaccinee antibody responses to MPXV A5, A27, A29 and VACV A27 were not significantly different from the negative samples (with the exception of MPXV A5, 63 days PD2) when compared by Dunnett’s multiple comparisons test. An increase in antibodies was observed for all other antigens 24 days PD1. For these eight antigens, antibodies were seen to be significantly boosted 14 days PD2 which waned at subsequent timepoints. Antibodies to MPXV E8, M1 and VACV B5 remained significantly higher than the negative samples up to the final timepoint of 240 days PD2. With some variation, antibodies to MPXV B6 remained significantly higher up to 122 days PD2, to MPXV A35 up to 43 days PD2, to MPXV B2 up to 185 days PD2, to VACV A33 up to 157 days PD2, and to MPXV H3 up to 157 days PD2. In contrast, antibody responses in post-MPXV infection samples were significantly higher for all antigens tested.

**Figure 5.**
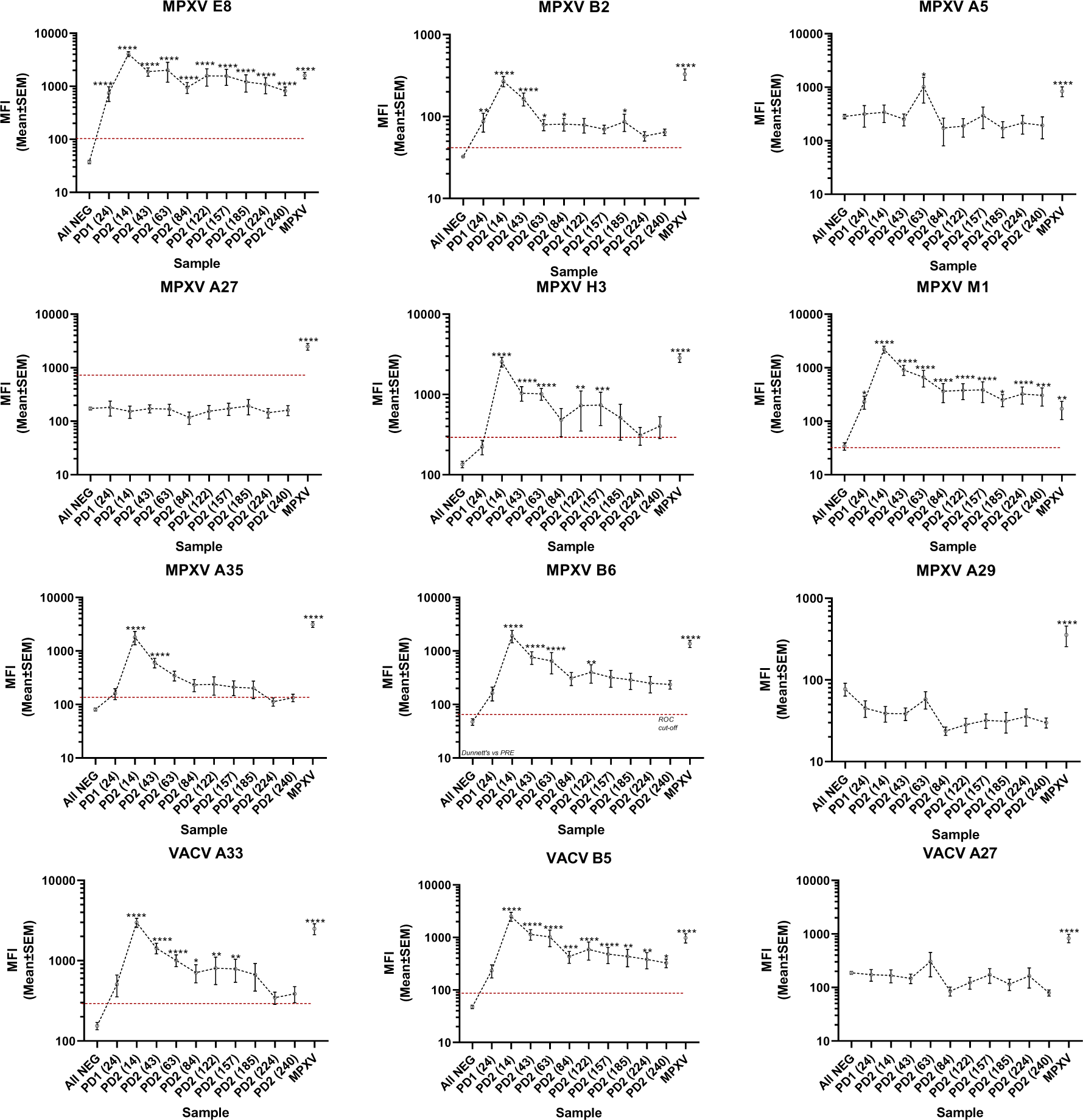
Longitudinal IgG Responses to Multiple MPXV and VACV Antigens Following IMVANEX Vaccination. MFIs of negative samples, post-MPXV infection samples and post-IMVANEX samples 24 days post one (PD1 (24)) and 14 to 240 days post two (PD2) doses to 12 MPXV and VACV antigens. Each graph represents the results for an individual antigen. Each point represents mean MFI ±SEM for all samples. Significance compared to the negative samples was calculated using Dunnett’s multiple comparisons test (* = <0.05; ** = <0.01; *** = <0.001; **** = <0.0001). Negative samples (n=435); post-MPXV infection samples (n = 24); PD1 (day 24, n = 7); PD2 (day 14, n = 8); PD2 (day 43, n = 10); PD2 (day 63, n = 7); PD2 (day 84, n = 7); PD2 (day 122, n = 7); PD2 (day 157, n = 7); PD2 (day 185, n = 6); PD2 (day 224, n = 8); PD2 (day 240, n = 8). The red dotted line represents the assay cut-off MFI where one is used.

Using the antigen MFI cut-offs and classification system as detailed previously, each sample was scored and classified as either negative, post-IMVANEX vaccination or post-MPXV infection (**Figure 6**). Of the 435 negative samples tested, 428 (98.4%) were correctly classified negative, six (1.4%) were incorrectly classified as post-vaccination and one (0.2%) was incorrectly classified as post-MPXV infection (data not shown). Of the 24 post-MPXV infection samples tested, 21 (87.5%) were correctly classified post-MPXV infection tested, two (8.3%) were incorrectly classified as negative and one (4.2%) was incorrectly classified as post-vaccination. Of the 75 post-vaccination samples (taken 24 days PD1 and between 14 to 240 days PD2) tested, 68 (90.7%) were correctly classified as post-vaccination and 7 (9.3%) were incorrectly classified as negative. Of those seven post-vaccination samples incorrectly classified as negative, one was taken at 24 days PD1, one was taken 122 days PD2, one was taken 185 days PD2, three were taken 220 days PD2 and one was taken 240 days PD2.

**Figure 6.**
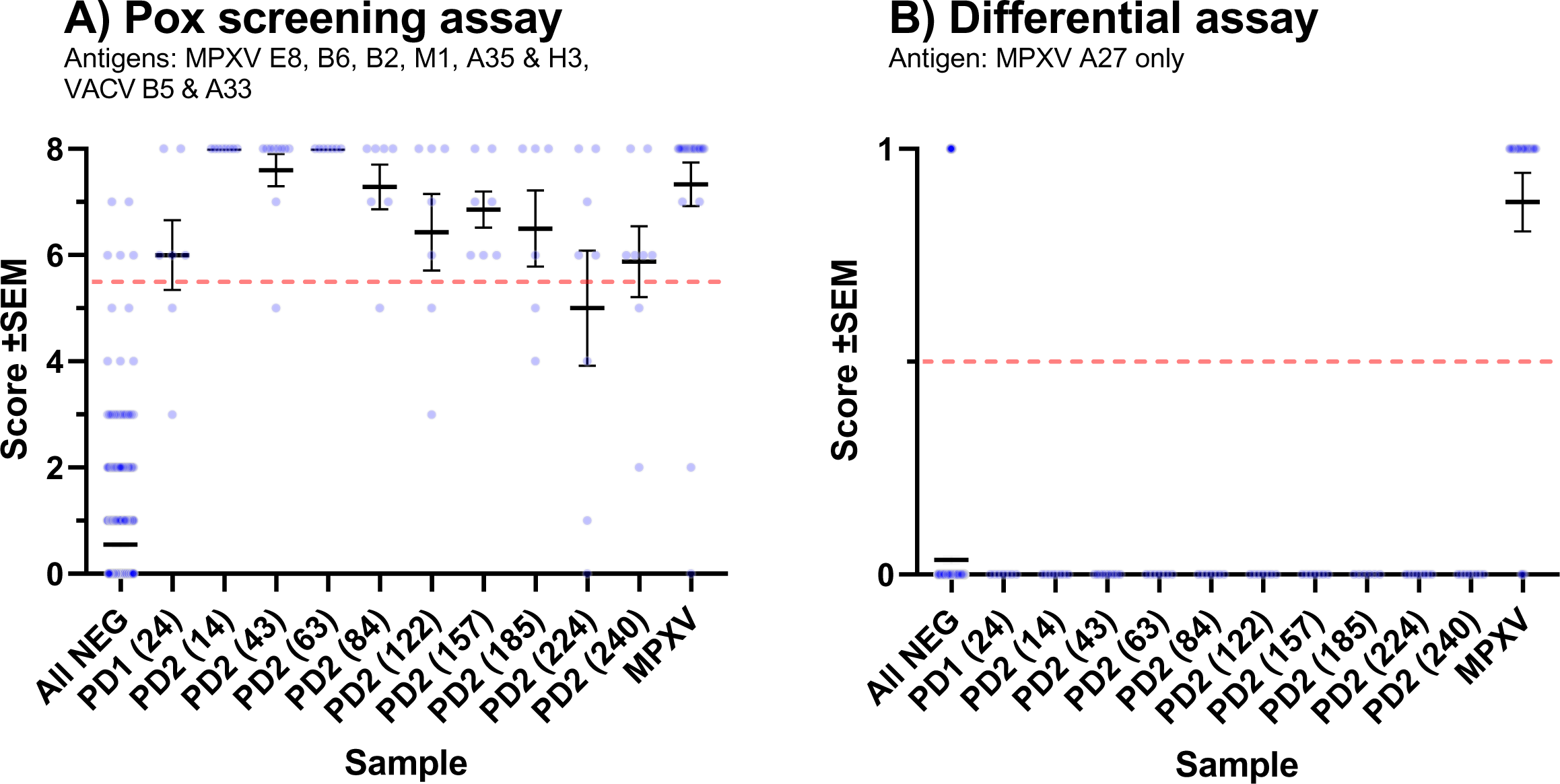
Positivity Scores of Post-IMVANEX Vaccination and Post-MPXV Infection Samples. Multiplexed Mpox assay positivity scores of negative samples, post-MPXV infection samples and post-IMVANEX samples 24 days post one (PD1 (24)) and 14 to 240 days post two (PD2) doses. (**A – Pox screening assay**) Samples with an MFI above the cut offs for VACV B5, A33, and MPXV E8, B6, B2, M1, A35 and H3 were given a score out of 8 (one for each antigen). (**B – Differential assay**) Samples with an MFI above the cut-off for MPXV A27L received a positivity score of 1. Each point represents the positivity score on an individual sample. Each solid line represents mean positivity score ±SEM. Negative samples (n=435); post-MPXV infection samples (n=24); PD1 (day 24, n=7); PD2 (day 14, n=8); PD2 (day 43, n=10); PD2 (day 63, n=7); PD2 (day 84, n=7); PD2 (day 122, n=7); PD2 (day 157, n=7); PD2 (day 185, n=6); PD2 (day 240, n=8); PD2 (day 240, n=8). The red dotted line represents the positivity score cut-off.

## Discussion

We have developed a bead-based multiplexed assay capable of measuring IgG responses to twelve Orthopoxvirus proteins. The antibody response to a combination of eight of these MPXV and VACV proteins (MPXV E8, B6, B2, M1, A35 and H3, and VACV B5 and A33) was able to distinguish positive samples, either post-vaccination or infection, from negative samples (Pox screening assay) and the IgG response to MPXV A27 could accurately distinguish post-MPXV infection from negative and post-vaccination samples (Differential assay). IgG responses to VACV A27, MPXV A29 and MPXV A5 provided little advantage in distinguishing between infection or vaccination despite significantly higher reactivity to these antigens measured in post-MPXV infection sera compared to vaccinated individuals. These 12 antigens were selected from this assay due to their immunodominance in response to MPXV infection and smallpox vaccination as described previously [44, 45].

There are two predominant infectious forms of MPXV, known as the mature virion (MV) and the enveloped virion (EV), each expressing different proteins on their surface membranes [53]. For example, MPXV proteins A29, E8, H3, and M1 are expressed on the MV surface membrane whilst A35, B2 and B6 are expressed on the membrane of the EV [54]. As such, when assessing the immunogenicity of vaccination, individual neutralisation assays must be performed using preparations of either MV or EV to include all relevant antigens. Assays, such as the multiplexed Mpox Luminex assay described here, can measure both MV and EV associated antigens at once far more conveniently and we are currently assessing the correlation of individual IgG responses to these antigens to both MPXV and VACV neutralisation.

### Differences Between IgG Responses to Vaccination and Infection

IgG responses to MPXV protein M1 were far more pronounced in those receiving smallpox vaccination when compared to convalescent MPXV infection samples (**Figure 1**). Despite this difference, responses were not sufficiently distinct to differentiate post-MPXV infection samples from post-smallpox vaccination (**Figure 2C**). Limited antibody responses to MPXV M1 protein post-MPXV infection has been previously reported and differences in expression levels between MPXV and VACV is suspected in causing this contrasting immunity [45, 46, 55]. The inability of MVA-BN to replicate in the host or protein changes between MPXV and VACV may also be responsible. However, the MPXV M1 protein is high conserved in all Orthopoxviruses with a sequence identity of 98.8% between MPXV and VACV [40]. MPXV M1, or L1 in VACV, is a surface membrane protein expressed on intracellular mature virus (IMV) and part of the entry/fusion complex (EFC) involved in cell entry and membrane fusion [56]. With MPXV M1 identified as an important immunogen, capable of producing potent neutralising antibodies, further work is needed to understand this difference in responses to VACV and MPXV and how this plays a role in immunity induced by infection or vaccination with Orthopoxviruses [36, 37, 49].

The sequence identity of the MPXV and VACV antigen homologues used in this study are high with MPXV B6R sharing 95.9% sequence identity with VACV B5R, MPXV A35R sharing 96.1% sequence identity with VACV A33R and MPXV A29L sharing 93.6% sequence identity with VACV A27L [40]. Despite this homology, antibody response to smallpox vaccination showed preferential binding to VACV antigens over their MPXV counterparts with the reverse being true for convalescent sera post-MPXV infection (**Figure 4**). This difference was most clear for MPXV A35 and VACV A33 and mirrors the findings of the singleplex Mpox ELISA [45], further demonstrated in this analysis through the wider dynamic range of the described assay. The specificity of this response is additionally supported by work showing that naïve individuals receiving two doses of the MVA-BN vaccine produce much lower neutralising titres against MPXV when compared to those either previously infected with MPXV or immunised with a second generation smallpox vaccine [57]. The reasons for and consequences of this stark difference require further investigation and, whilst smallpox vaccines provide cross-protection against MPXV, this suggests MPXV specific vaccines may improve protection against disease and reduce vaccine breakthroughs [58].

Antibody responses to these antigens following infection with MPXV clade I were not investigated in this study die to limited access to sera. With the ongoing outbreak in the DRC, whether the Multiplexed Mpox Luminex assay can be used to assess antibodies to this clade of MPXV is of importance.

### Singleplex Mpox ELISA Versus Multiplexed Mpox Luminex Assay

Comparison of the results from the singleplex Mpox ELISAs and multiplexed Mpox Luminex assay showed a good correlation for most of the antigens assessed (**Figure 3**). However, the correlations of the two assays for antigens MPXV A5, A29 and H3 was poor. For MPXV A5, this was driven primarily by higher results in the ELISA for all sample types compared to the Luminex assay whereas the reverse was true for MPVX A29 and MPXV H3. Interestingly, both post-MPXV infection and post-vaccination samples were higher in the Luminex assay for MPXV H3 whilst only post-MPXV infection samples were higher in the Luminex assay for MPXV A29. In ELISAs, antigens are attached to the assay plate via passive absorption through hydrophobic and hydrophilic interactions whereas antigens are covalently linked to assay beads when using the Luminex platform via EDC. Differences in these methods and the properties of the antigens may account for this difference. Although not unique in the proteins used, MPXV A29 and MPXV H3 are comparably more basic, and contain more hydrophobic and fewer neutral residues than the other antigens (**Table S2**). This may have an impact on absorption to the ELISA plates and levels of denaturation.

The relationship between ELISA OD_450nm_ and Luminex MFI was hyperbolic with samples reaching the signal limit of the ELISA assay spreading out in the Luminex assay (**Figure 3** and **Figure S2**). This difference in the assays demonstrates the wider dynamic range of the Luminex platform over ELISAs. The ability of the Luminex platform to measure multiple antigens simultaneously provides additional benefits over the singleplex Mpox ELISA, saving time and resources whilst providing richer information at comparable levels of sensitivity and specificity [45]. Both the singleplex Mpox ELISAs and multiplexed Mpox Luminex assay performed similarly in their abilities to identify positive samples and to differentiate vaccinated samples from convalescent samples with sensitivities and specificities ranging from 87.5% to 98.9%, respectively. It is worth noting that the two PCR-confirmed Mpox samples that returned negative results in the Luminex assay were also negative when run through the ELISAs. Additionally, the ease at which antigens can be removed or added to the Luminex assay provides versatility over other multiplexed platforms, such as MSD [59].

### Longitudinal IgG Response to IMVANEX Vaccination

IgG antibodies to twelve MPXV and VACV antigens following vaccination with IMVANEX were assessed using the multiplexed Mpox Luminex assay (**Figure 5**). Antibodies were measured 24 days after one dose of the vaccine and at multiple timepoints between 14 to 240 days after a second dose. In contrast to post-MPXV infection sera, no antibodies to MPXV A5, A27, A29 and VACV A27 antigens were detected post-vaccination with IMVANEX. As previously stated, differences in the expression levels of these proteins between MPXV Clade II(b) and MVA-BN and inability of MVA-BN to replicate in the host may be responsible for the lack of antibodies to these antigens post-IMVANEX vaccination. MPXV A29 protein and its orthologue in VACV (A27) mediate the interaction of the MV with cell surface heparan and have been shown to induce neutralising antibodies against the virus [60, 61]. Similar muted responses to VACV A27 have also been noted following Dryvax, ACAM200 and JYNNEOS vaccination with much higher responses post-Lister strain derived MVA vaccination or MPXV infection [41, 43, 46, 61]. Identifying why the antibody response is so low in individuals receiving some smallpox vaccines may help improve the immunogenicity of the next generation of vaccines.

All other antigens assessed produced measurable IgG responses after one-dose which was subsequently boosted after the second dose. Aside from the poxin MPXV B2, each of these proteins have previously been associated with producing neutralising antibodies [37, 46, 47]. High levels of antibodies to MPXV M1 and VACV B5 were maintained up to the last timepoint measured whilst antibodies to the remaining antigens had waned to levels comparable to 24 days post-one dose of IMVANEX. The impact of this waning on neutralisation titres was not assessed but warrants further investigation to aid in the understanding of the longevity of protection provided by IMVANEX against MPXV infection, and if correlated, this assay may offer a simple and rapid solution for assessing immunity against MPXV.

## Conclusions

With additional benefits over current serological MPXV assays, we believe this assay provides substantial insight to the current global outbreak of Mpox, facilitating with our understanding of the immunology elicited by both MPXV-infection and Smallpox-vaccination. One specific advantage over other current assays is the ability to differentiate responses to infection and vaccination using MPXV A27 [62].We are now applying these assays to a number of other cohorts, such as longitudinal post-MPXV infection samples and serosurveillance studies to further validate the assay and to understand which immunological markers are useful for serosurveillance. We also hope to utilise the dual reporter capability of the Luminex xMAP INTELLIFLEX® system to assess IgM responses to infection and vaccination alongside IgG. Correlation of results from this assay with current neutralisation assays will also provide important insights into which antibodies are necessary for protection. Furthermore, with the generation of MPXV-specific vaccine candidates, this assay offers a versatile method for assessing vaccine immunogenicity and is highly amenable to the addition of new antigens.

## Data Availability

All data produced in the present study are available upon reasonable request to the authors

## Supplementary

**Figure S1.**
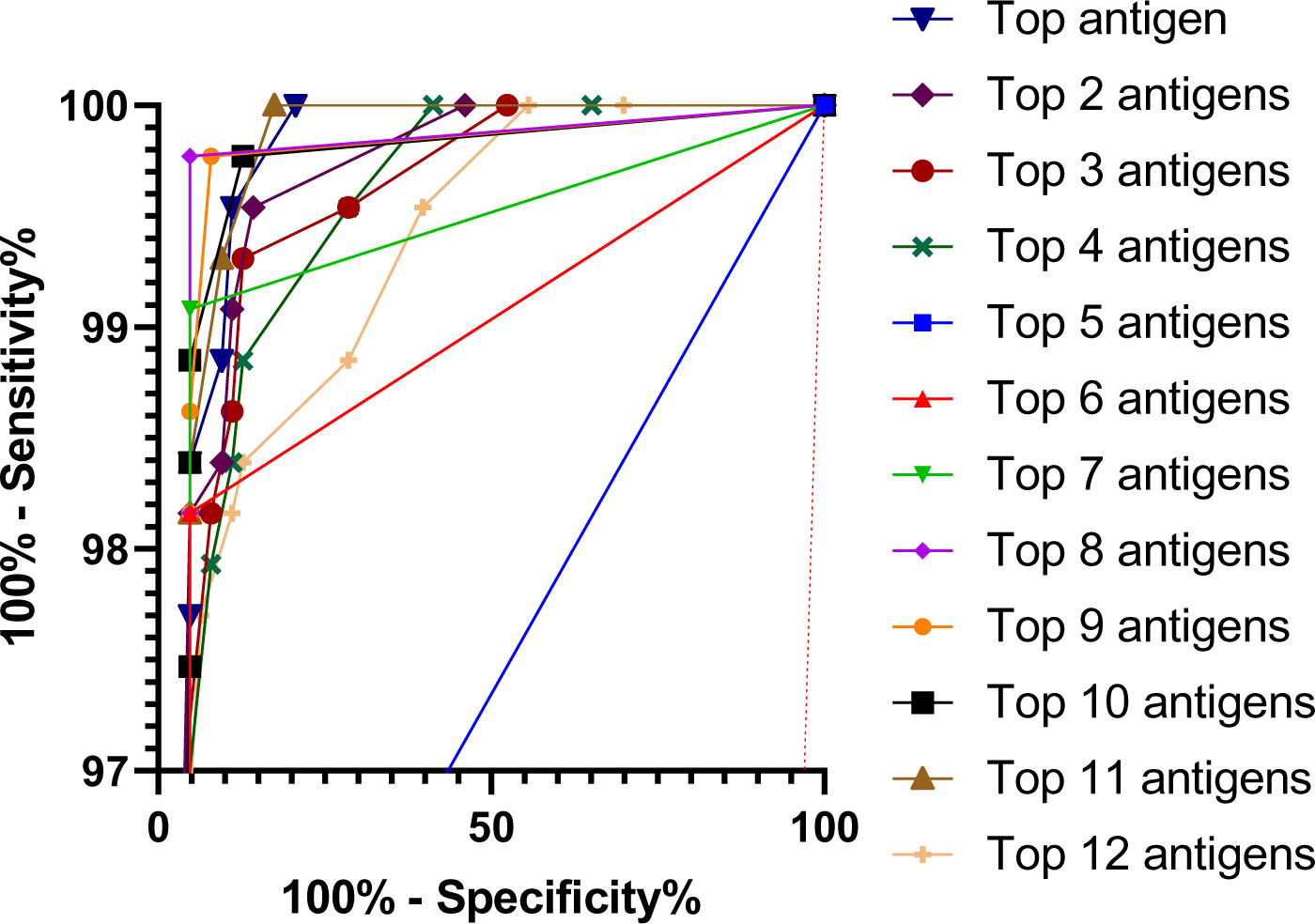
Antigen Combination ROC Curves for Positivity Scores (Negative Versus Positive Samples) ROC curves comparing the positivity scores of negative and positive samples for each combination of antigens. Positivity scores were calculated based on how many antigen cut-offs a sample was above (one for each antigen). The order of antigens was the following: VACV B5 (top), MPXV E8, B6, B2, VACV A33, MPXV M1, A35, H3, A29, A5, A27 and VACV A27 (bottom). The characteristics of each ROC curve and optimal cut-offs are detailed in **Table 1**. Each solid line represents the ROC curve for each antigen combination. Dotted red line represents the random classifier (i.e., sensitivity = specificity). Negative samples (n= 435) and positive samples (n=24 post-MPXV infection and n=39 post-IMVANEX vaccination (14 to 144 days post-two doses)).

**Table S1.** Samples Used in This Study. Details and numbers of samples used in this study. Median ages (years) and median times since vaccination/infection (days) are shown. CMV – cytomegalovirus. EBV – Epstein Barr virus. RF – Rheumatoid factor. VZV – Varicella zoster virus.

| Description | Number of samples | Median age (yrs) | Median time since vaccination/infection |
| --- | --- | --- | --- |
| Negatives |  |  |  |
| Negative Confounder – CMV | 52 | 34 | N/A |
| Negative Confounder – EBV | 34 | 25 | N/A |
| Negative Confounder – RF positive | 63 | 15 | N/A |
| Negative Confounder – VZV | 44 | 32 | N/A |
| Pre-IMVANEX Vaccination | 18 | 27 | N/A |
| Paediatric Negatives | 224 | 3 | N/A |
| Post-MPXV Infection |  |  |  |
| Convalescent Mpox (Clade II(a)) | 2 | 37 | 7 days |
| Convalescent Mpox (Clade II(b)) | 22 |  | 81 days |
| Post-IMVANEX Vaccination |  |  |  |
| IMVANEX post-dose 1 | 7 | 26 | 24 days |
| IMVANEX post-dose 2.1 | 8 | 26 | 14 days |
| IMVANEX post-dose 2.2 | 10 | 26 | 43 days |
| IMVANEX post-dose 2.3 | 7 | 26 | 63 days |
| IMVANEX post-dose 2.4 | 7 | 26 | 84 days |
| IMVANEX post-dose 2.5 | 7 | 26 | 122 days |
| IMVANEX post-dose 2.6 | 7 | 26 | 157 days |
| IMVANEX post-dose 2.7 | 6 | 26 | 185 days |
| IMVANEX post-dose 2.8 | 8 | 26 | 224 days |
| IMVANEX post-dose 2.8 | 8 | 26 | 240 days |

**Table S2.** Properties of Antigens Used in this Study. Percentage hydrophobic, acidic, basic, and neutral residue composition of the antigens used in this study. The overall protein charge, pI and size (kDa) of each protein are also shown. Antigen properties were calculated using protein sequences provided by each manufacturer with protein isoelectric point and hydrophobicity/hydrophilicity calculators [63, 64].

| Antigen | Product code | Residues |  |  |  | Attribute | pI | kDa |
| --- | --- | --- | --- | --- | --- | --- | --- | --- |
|  |  | Hydrophobic | Acidic | Basic | Neutral |  |  |  |
| MPXV antigens |  |  |  |  |  |  |  |  |
| A35 | 40886V08H | 32.56% | 12.40% | 10.85% | 44.19% | Acidic | 4.58 | 13.74 |
| B6 | PXP6031 | 29.62% | 14.23% | 8.85% | 47.31% | Acidic | 4.39 | 28.82 |
| B2 | ABX620113 | 28.52% | 18.36% | 6.25% | 46.88% | Acidic | 3.75 | 27.91 |
| A27 | PXP6054 | 25.00% | 26.12% | 20.52% | 28.36% | Acidic | 4.79 | 31.79 |
| E8 | PXP6055 | 38.54% | 10.42% | 13.54% | 37.50% | Basic | 6.57 | 31.63 |
| M1 | 40904V07H | 38.33% | 8.89% | 8.33% | 44.44% | Acidic | 5.28 | 19.32 |
| A5 | REC32033 | 37.72% | 10.68% | 8.90% | 42.70% | Acidic | 4.81 | 30.97 |
| A29 | 40891V08E | 32.94% | 16.47% | 21.18% | 29.41% | Basic | 8.14 | 9.72 |
| H3 | 40893V08H1 | 45.68% | 13.89% | 13.89% | 26.54% | Neutral | 5.24 | 32.71 |
| VACV antigens |  |  |  |  |  |  |  |  |
| B5 | 40900V08H | 30.00% | 13.46% | 8.46% | 48.08% | Acidic | 4.42 | 28.83 |
| A33 | 40896V07E | 32.56% | 12.40% | 10.08% | 44.96% | Acidic | 4.67 | 14.23 |
| A27 | 40897V07H | 36.36% | 20.91% | 19.09% | 23.64% | Acidic | 5.2 | 12.62 |

**Table S3.** Mean MFIs of Negative, Post-Infection and Post-IMVANEX Samples. Mean MFIs of post-MPXV infection (n=24), post-IMVANEX vaccination (timepoints between 14 to 144 days post-two doses; n=39) and negative samples (pre-IMVANEX vaccination: n=18; paediatric negatives: n=224 and negative confounders: n=193) to each of the 12 MPXV and VACV antigens tested. P values of post-MPXV infection and post-IMVANEX vaccination samples versus negative samples MFIs were calculated using Dunnette’s multiple comparisons test.

| *Antigen* | **Mean MFIs** |  |  | **P value vs. Negatives** |  |
| --- | --- | --- | --- | --- | --- |
|  | *Negatives* | *Post-Infection* | *Post-IMVANEX* | *Post-Infection* | *Post-IMVANEX* |
| *MPXV Antigens* |  |  |  |  |  |
| B6 | 47.43 | 1366 | 832.4 | <0.0001 | <0.0001 |
| E8 | 37.47 | 1601 | 2129 | <0.0001 | <0.0001 |
| A35 | 80.77 | 3127 | 672.3 | <0.0001 | <0.0001 |
| B2 | 32.44 | 329.5 | 140.4 | <0.0001 | <0.0001 |
| A5 | 284.6 | 840 | 380.8 | <0.0001 | 0.5649 |
| A27 | 173.3 | 2488 | 154.4 | <0.0001 | 0.9571 |
| M1 | 34.21 | 171.6 | 937.6 | 0.0388 | <0.0001 |
| H3 | 134.5 | 2855 | 1189 | <0.0001 | <0.0001 |
| A29 | 77.22 | 356.1 | 37.51 | <0.0001 | 0.6549 |
| *VACV Antigens* |  |  |  |  |  |
| A33 | 154.1 | 2493 | 1432 | <0.0001 | <0.0001 |
| B5 | 47.54 | 978.2 | 1174 | <0.0001 | <0.0001 |
| A27 | 187.4 | 829.7 | 164.2 | <0.0001 | 0.8502 |

